# The Benefits and Harms of Open Notes in Mental Health: A Delphi Survey of International Experts

**DOI:** 10.1101/2021.04.21.21255785

**Authors:** Charlotte Blease, Anna Kharko, Maria Hagglund, Stephen O’Neill, Deborah Wachenheim, Liz Salmi, Kendall Harcourt, Cosima Locher, Catherine M. DesRoches, John Torous

**Author notes:** **Corresponding author:** C. Blease, Ph.D., General Medical and Primary Care Research, Beth Israel Deaconess Medical Center, 330 Brookline Avenue, Boston, MA 02215, Massachusetts, USA.

## Abstract

**Importance:** As of April 5, 2021, as part of the 21^st^ Century Cures Act, new federal rules in the U.S. mandate that providers offer patients access to their online clinical records.

**Objective:** To solicit the view of an international panel of experts on the effects on mental health patients, including possible benefits and harms, of accessing their clinical notes.

**Design:** An online 3-round Delphi poll.

**Setting:** In round 1 open-ended questions solicited feedback on the benefits and harms to patients of reading their mental health notes. Responses were coded to produce itemized statements. In Round 2 participants were asked to rate their agreement with each item along 7-point Likert scales. Responses were analyzed for consensus, set at a predetermined interquartile range of ≤ 1. In Round 3 items that did not reach consensus were redistributed.

**Participants:** International experts identified as clinicians, chief medical information officers, patient advocates, and informaticians with extensive experience and/or research knowledge about patient access to mental health notes.

**Main Outcomes, and Measures:** An expert-generated consensus on the benefits and risks of sharing mental health notes with patients.

**Results:** A total of 70 of 92 (76%) experts from 6 countries responded to Round 1. A qualitative review of responses yielded 88 distinct items. A total of 56 of 70 (80%) experts responded to Round 2, and 52 of 56 (93%) responded to Round 3. Consensus was reached on 65 of 88 (74%) of survey items.

**Conclusions and Relevance:** This iterative process of survey responses and ratings yielded consensus that there would be multiple benefits and few harms to patients from accessing their mental health notes. Questions remain about the impact of open notes on professional autonomy, and further empirical work into this practice innovation is warranted.

## Introduction

As of April 5, 2021, new federal rules in the U.S. mandate that all health providers offer patients online access to their clinical notes.[1,2] These new information sharing rules resulting from the 21st Century Cures Act of 2016 require rapid and full online access to test results, medication lists, referral information, and progress notes (so-called ‘open notes’). Psychotherapy notes are exempt, and “information blocking” is permitted, if doing so “…will substantially reduce the risk of harm” where this is understood as physical harm to a patient or to another person or if there is a privacy exception (§ 171.201(a) p. 704).[1] Licensed health professionals can decide what constitutes a substantial risk when working “…in the context of a current or prior clinician-patient relationship” (p. 702).

Sharing access to mental health notes is controversial, and in countries that have already opened notes to patients, many psychiatric organizations have resisted implementing the practice.[3] In surveys mental health clinicians worry that patients will become anxious, confused or upset by what they read; many also anticipate access will lead to increased work burdens.[4–6] Findings drawn predominantly from primary care suggest that mental health patients may derive benefits from accessing their clinical notes including feeling more in control of their care, better remembering their care plan, and better understanding the rationale for medications.[7,8] However, sharing mental health notes could present more challenges than in other clinical specialties. Currently, only a few pilot surveys have examined patients’ experiences in specialized mental health settings, and while the results are encouraging, at least some patients reported feeling disrespected or judged by what they read.[9–12] In existing studies, small sample sizes, the exclusion of patients with serious mental illnesses (SMI) or personality disorders, and the possibility of clinician and patient response biases also limit the potential for informative inferences about psychiatric care.[13]

In light of limitations, clinicians may be uncertain about when it is appropriate to hide information. For example, it is unclear whether access may exacerbate harms among mental health patients. On the other hand, blocking information may deny patients important opportunities to better understand, manage, and become engaged in their care. Considering the pressing need for greater clarity in light of the new U.S. healthcare regulations taking effect in April 2021, and while awaiting further empirical research to inform best practice, our goal was to establish expert-led consensus about the potential benefits and harms of sharing notes in mental healthcare including among persons with serious and complex needs. Therefore, we set out to conduct the first Delphi poll in the field, encompassing a range of stakeholders including patient advocates.[14]

## Methods

### Background

Delphi polls are a recognized methodology for exploring the consensus views of experts in an emerging field, providing more accurate predictions than other forecasting techniques.[15–17] The approach is designed to pool the views of a purposive sample of identified experts who are invited to answer a series of open-ended questions on a focused topic. Participants are next required to reassess their initial judgements, in light of aggregate group feedback, until consensus is obtained. While some Delphi polls employ the use of face-to-face meetings to help establish areas of agreement, more robust approaches employ anonymous, iterative techniques to avoid the influences of group think or dominant personalities.[16,18] Delphi polls are particularly well-suited to gauging expert consensus and have been used extensively in healthcare policy and management, including in psychiatry.[16,19,20]

### Approach

We used a modified Delphi technique structured around three discrete online surveys.[21,22] In Round 1, questions are open-ended requiring free-text responses which are then aggregated and translated into survey items. In Round 2, these experts are provided with the list of survey items and asked to rate their level of agreement using Likert scales. To reduce survey fatigue, those statements that reach a predefined level of consensus are omitted for Round 3, and remaining items are recirculated. In this final round, participants are reminded of their previous judgment of each of the remaining items and are also furnished with the median response of the other experts and invited to preserve or revise their previous response. While there is no universal agreement about the sample size for Delphi polls, a key objective of Delphi is to maximize the response rate between each round which has been demonstrated to improve the accuracy of consensus opinions and forecasts.[16,18,21]

### The expert panel

Using purposive sampling, the research team compiled a list of prospective participants with expertise on sharing mental health notes. We defined expertise as individuals with experience as: clinicians of sharing mental health notes with patients; patients with mental health diagnoses, including patient advocates with lived experience or knowledge of the practice; chief medical information officers or directors of divisions of health organizations that offered open notes to mental health patients; and/or, academic informaticians, who had published significant contributions within the field health informatics and patient access to clinical records. The research team was mindful to ensure gender, age, race, and international diversity, and specifically strove to include representation from countries and health systems where open notes have been implemented. To ensure a diversity of representation we also employed a snowballing technique by inviting chief medical information officers of health organizations that had opened psychiatric notes to assist us by providing names and contact details of mental health clinicians with experience of the practice. The final list of experts included 92 prospective participants. The study received approval from the Beth Israel Deaconess Medical Center Institutional Review Board in April 2020 (Protocol # 2020P000218) and the University of Plymouth, UK (Protocol # 19/20-1331). Invited participants were advised that the survey was voluntary and unpaid, that their responses would be anonymous to other participants, and that their identity was restricted to members of the research team. Identified experts were contacted via email in August 2020 with an invitation and link to the web-based survey. Invitees were informed that an adequate response time would be given between rounds and that they could choose to withdraw at any time. All respondents gave informed consent before participating.

### The Questionnaire

An electronic survey was created using Qualtrics (Qualtrics, Provo, UT). Since all prospective participants were fluent English speakers, the survey was only administered in this language. The poll incorporated the three-step modified Delphi technique, and data collection ran from August to November 2020. Participants were sent up to 4 reminders, 1 week apart, and given 4 to 5 weeks to respond to each round (see Figure 1). The first round comprised demographic questions and requested information about the nature of participants’ expertise with open notes. This was followed by six open-ended questions on sharing mental health notes plus one additional open-ended question allowing participants to comment on the survey or submit additional responses (see Appendix 1). The present Delphi study was based on respondents’ answers to two questions on the potential benefits and harms to patients of reading mental health notes. Participants were asked: “What, in your opinion, are the benefits – if any – of sharing mental health notes with patients?” and “What, in your opinion are the harms – if any – of sharing mental health notes with patients?” Answers to other questions formed the basis of a separate qualitative study that has been published elsewhere.

Following the closure of Round 1, descriptive content analysis was used to transform responses into lists of statements.[23] Coding was conducted by CB and independently reviewed by JT and MH and subsequent refinements were made. Comments that were unclear or deemed irrelevant to the question were excluded. Any disagreements in coding were settled by consensus among CB, JT, and MH. Statements were then transformed into survey items, and where possible participants’ exact phrasing was preserved. Survey items relating to benefits, and harms of sharing mental health notes formed two separate sections of the survey circulated in Round 2, and participants were requested to respond to each question using a variety of predefined 7-point Likert scales: 1 = *strongly disagree*, 2 = *moderately disagree*, 3 = *slightly disagree*, 4 = *neutral*, 5 = *slightly agree*, 6 = *moderately agree*, 7 = *strongly agree* (see Appendix 2). Prior to analyzing responses to Rounds 2 and 3, consensus to items was set at an interquartile range of ≤ 1.[18,24] After analysis of Round 2 responses, items that did not reach consensus were recirculated in Round 3. In Round 3, each participant received a bespoke survey in which they were reminded of their responses to items in Round 2 and provided with the median response of all participants. After competing closed-ended survey questions in each round, participants were also invited to provide any additional feedback. All survey items that reached consensus were collated into themes, and subthemes by CB and JT.

## Results

### Overview

A total of 70 of 92 (76%) participants from 6 countries responded to Round 1. Among respondents, 35 of 70 (50%) identified as male, and the mean age was 50 (see Table 1). Forty-seven of 70 (67%) reported currently working in clinical practice, and 7 of 70 (10%) reported lived experience of illness. The mean number of years of experience working as a clinician, an open notes researcher, or as a patient advocate was 16 years. All respondents left comments (5,760 words) which were typically brief (1 or 2 sentences). Replies were shorter when discussing benefits (2,639 words) than harms (3,121 words). As a result of descriptive analysis of free-text responses, the survey was expanded into 88 items, 42 items on the potential benefits of sharing mental health notes with patients and 46 on potential harms (see Appendix 2).

**Table 1.** Characteristics of Survey Respondents by Round

| **Characteristic** | **No. (%) of Respondents** |  |  |
| --- | --- | --- | --- |
|  | **Round 1**<br>**(n = 70)** | **Round 2**<br>**(n = 56 of 70**<br>**[80.0%])** | **Round 3**<br>**(n = 52 of 56**<br>**[92.8%])** |
| Gender |  |  |  |
| Male <sup>a</sup> | 35 (50) | 28 (50) | 26 (50) |
| Age range, y |  |  |  |
| 20 – 29 | 1 (1) | - | - |
| 30 – 39 | 19 (27) | 17 (30) | 17 (33) |
| 40 – 49 | 14 (20) | 11 (20) | 10 (19) |
| 50 – 59 | 19 (27) | 15 (27) | 14 (27) |
| ≥ 60 | 17 (24) | 13 (23) | 11 (21) |
| Ethnicity |  |  |  |
| Asian | 6 (9) | 4 (7) | 4 (8) |
| Black / African / Caribbean | 1 (1) | 1 (2) | 1 (2) |
| White | 59 (84) | 49 (88) | 46 (88) |
| Other | 2 (3) | 1 (2) | - |
| Declined to answer | 2 (3) | 1 (2) | 1 (2) |
| Country of residence |  |  |  |
| Canada | 3 (4) | 2 (4) | 2 (4) |
| Estonia | 1 (1) | 1 (2) | 1 (2) |
| Norway | 3 (4) | 3 (5) | 3 (6) |
| Sweden | 12 (17) | 10 (18) | 10 (19) |
| UK | 4 (6) | 4 (7) | 4 (8) |
| USA | 47 (67) | 36 (64) | 32 (62) |
| Currently works in clinical practice | 45 (64) | 34 (61) | 31 (60) |
| Psychiatry | 15 (21) | 10 (18) | 9 (17) |
| Primary care | 10 (14) | 8 (14) | 8 (15) |
| Clinical psychology / Psychotherapy | 7 (10) | 6 (11) | 6 (12) |
| Psychiatric Nursing | 4 (6) | 4 (7) | 4 (8) |
| Paediatrics | 3 (4) | 3 (5) | 2 (4) |
| Social work | 3 (4) | 1 (2) | - |
| Palliative care / Home hospice | 2 (3) | 1 (2) | 1 (2) |
| Peer support | 2 (3) | 2 (4) | 2 (4) |
| Hospitalist | 1 (1) | 1 (2) | 1 (2) |
| Radiology | 1 (1) | 1 (2) | 1 (2) |
| Occupation or expertise related to health / open notes informatics research / patient advocacy <sup>c</sup> |  |  |  |
| Clinician | 46 (66) | 35 (63) | 32 (62) |
| Researcher | 25 (31) | 21 (38) | 21 (40) |
| Chief Information Officer / Portal Director / Medical Director | 14 (20) | 12 (21) | 9 (17) |
| Patient Advocate / Person with Lived Experience | 5 (7) | 4 (7) | 4 (8) |
| Social Worker | 1 (1) | 1 (2) | 1 (2) |

<sup>a</sup> Including one transgender male.
<sup>b</sup> Including Adult, Child and Adolescent Psychiatry
<sup>c</sup> Numbers & percentages may add up greater than the total n, as some participants reported multiple expertise.

In Round 2, 56 of 70 (80%) experts participated in the online survey and reached consensus on 22 of 88 items (25%). The mean age was 50 years old, and the mean number of years of experience working as a clinician, open notes researcher, or as a patient advocate was 17 years. In Round 3, 52 of 56 (93%) experts responded, reaching consensus on a further 43 items. In total, consensus was achieved for 65 of 88 items (74%). Among Round 3 respondents, the mean age was 49, and the mean number of years of experience working as a clinician, an open notes researcher, or as a patient advocate was 17 years. Table 2 presents the items that reached consensus after each round. Following thematic analysis, all items that reached consensus were categorized into four themes.

**Table 2.**
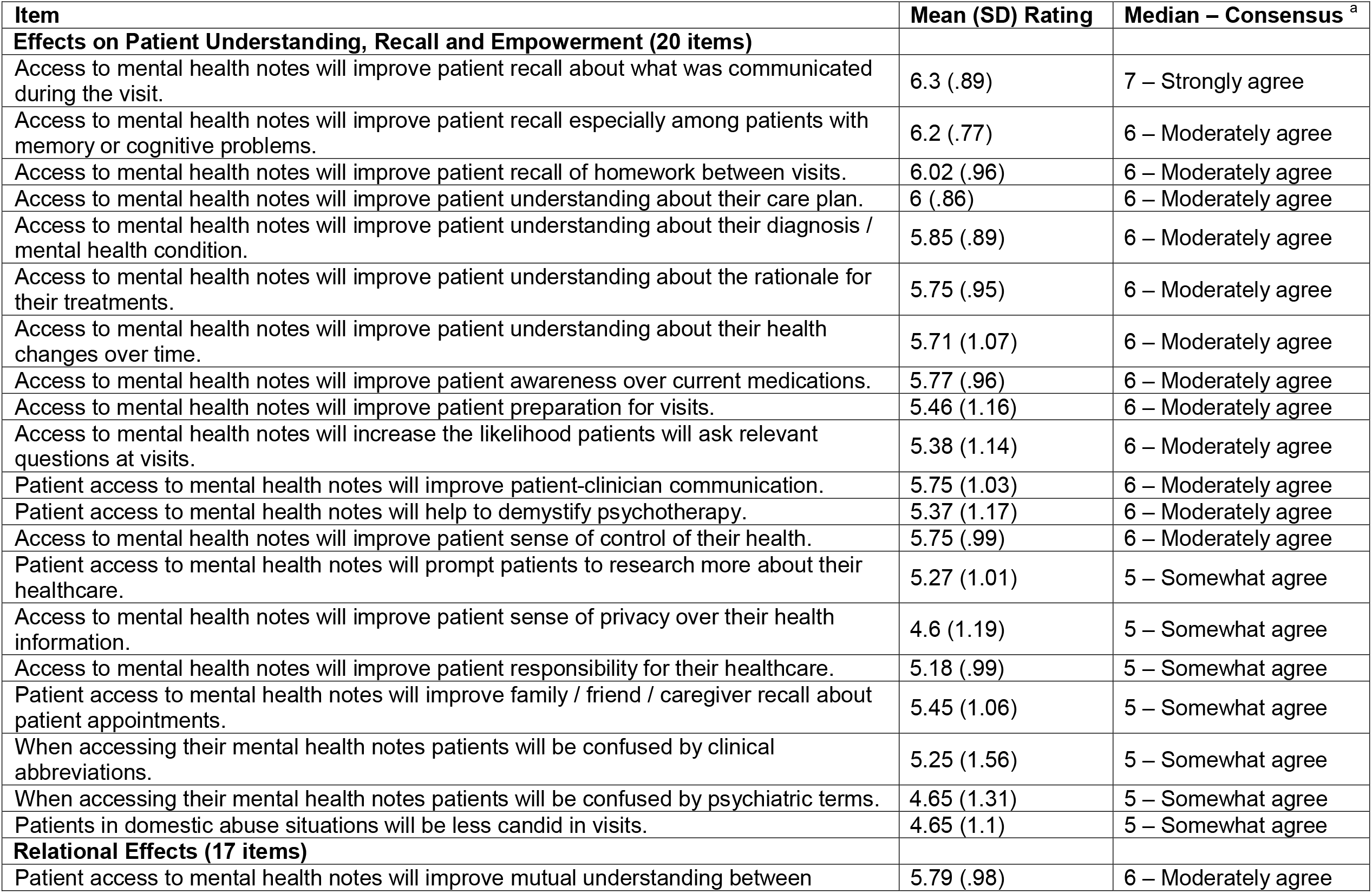
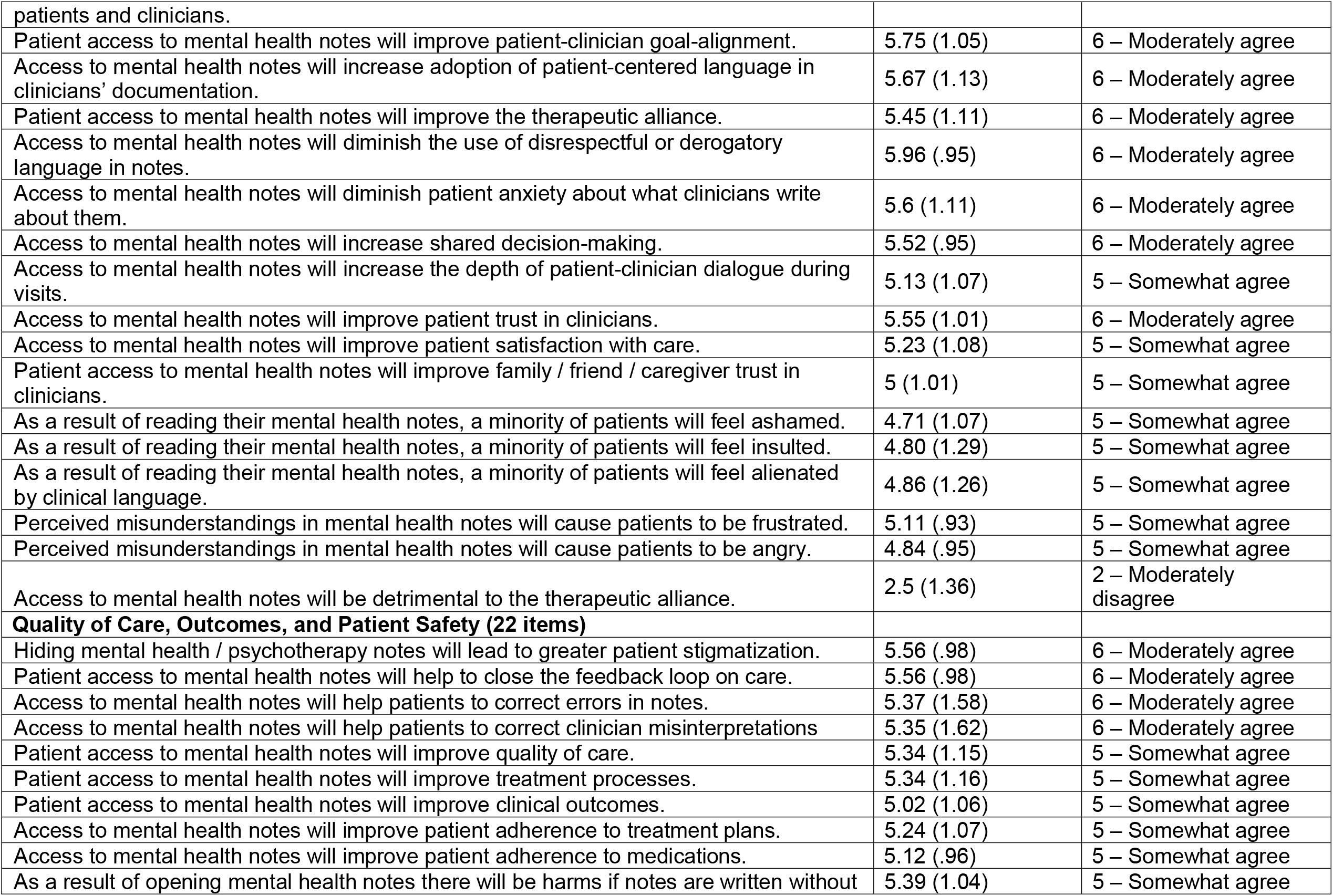
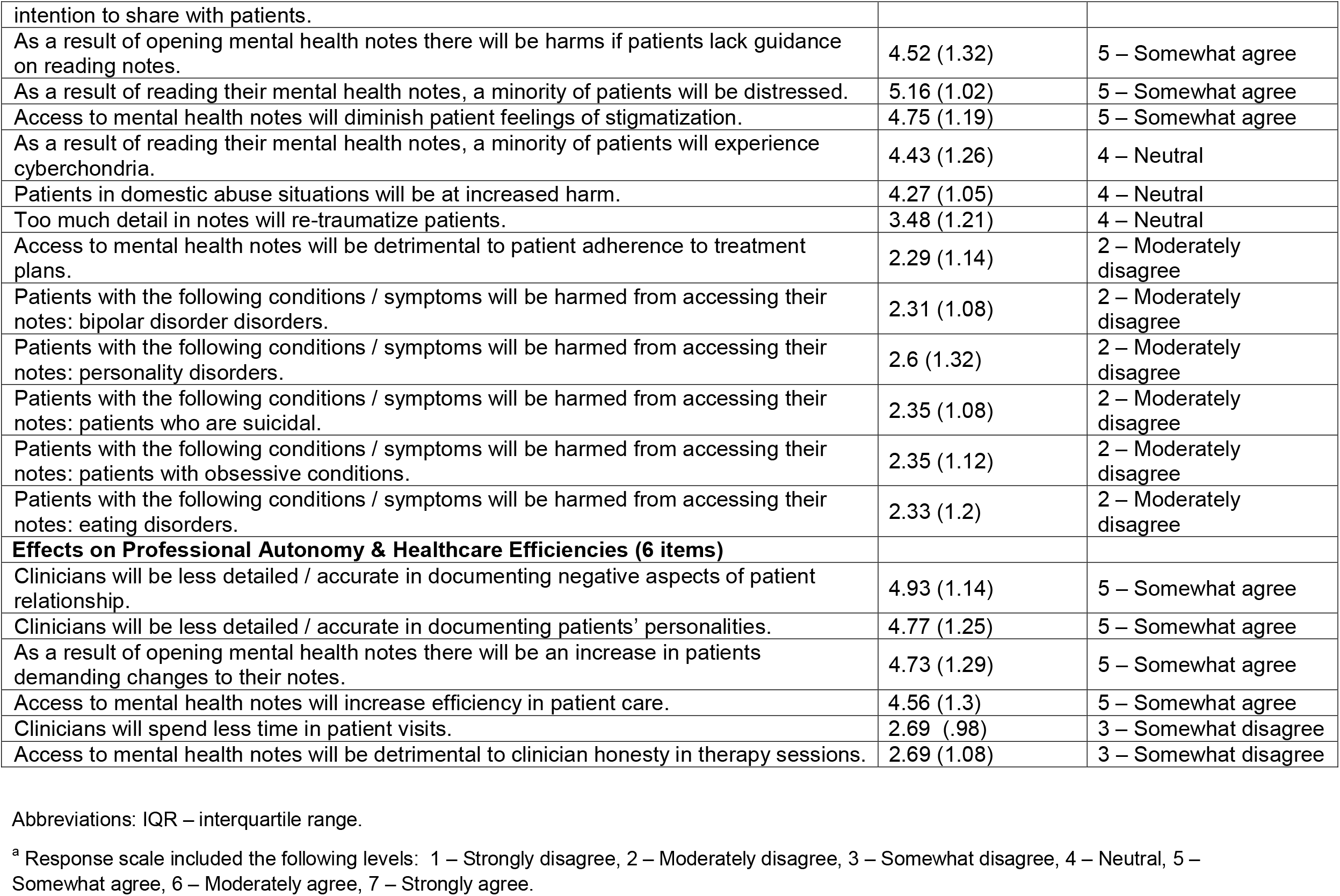
Items on the Benefits and Harms of Open Notes in Mental Healthcare that Reached Consensus

### Themes

#### Effects on Patient Understanding, Recall, and Empowerment

Panelists agreed that open notes could aid patient recall about what was communicated in visits, and that access to documentation could improve patient-clinician communication, including patient understanding about their diagnosis, prescribed medications, and the rationale for treatments. They also agreed that access could improve mental health patients’ sense of control over their healthcare, insight about their health, and help them better prepare for visits.

#### Relational Effects

Reflecting on whether access could affect the patient-clinician relationship, there was agreement among experts that open notes could improve trust, enhance the therapeutic alliance, increase patient-clinician goal-alignment, and strengthen shared decision-making. Panelists also believed that patient access would increase the adoption of patient-centered language and diminish the use of disrespectful or derogatory language in documentation. There was also agreement that access to clinical notes could diminish patients’ anxiety about what clinicians might write about them.

#### Quality of Care, Outcomes, and Patient Safety

Commenting on the potential for adverse effects, panelists *disagreed* that open notes would be harmful to patients with serious mental health diagnoses, specifically including persons with bipolar disorders, major depressive disorders, personality disorders, eating disorders, obsessive conditions, and individuals who are suicidal. Panelists agreed that access could help patients to correct errors in notes, including clinician misinterpretations, and that offering access could help to close the feedback loop on care. Furthermore, there was consensus that hiding mental health or psychotherapy notes might lead to greater patient stigmatization or harm. However, participants “somewhat agreed” that harms could arise if clinicians wrote notes without the intention that they might be read by patients, or if patients lacked guidance on how to read their notes. Panelists also “somewhat agreed” that access could improve treatment, including medication, adherence.

#### Effects on Professional Autonomy and Healthcare Efficiencies

Expert panelists “somewhat agreed” that as a result of access, there could be an increase in patients demanding changes to their clinical notes. Commenting on the effects on professional autonomy, panelists also “somewhat agreed” that mental health clinicians would be less detailed/accurate in documenting negative aspects of the patient relationship, details about patients’ personalities, or symptoms of paranoia in patients. However, experts predicted that patient access to mental health notes could increase efficiency in care delivery.

## Discussion

Currently, limited attention has been paid to the benefits and harms of reading mental health notes among patients with psychiatric diagnoses, including persons with SMI. In view of the lack of large-scale studies into the effects on mental health patients’ experiences, we used a rigorous modified Delphi approach to establish the consensus views about this practice innovation among an international stakeholder group of clinicians, patients, chief medical information officers, and informaticians. The final results comprise the first consensus-driven statement on the benefits and harms of online access to mental health notes.

In this Delphi poll there was consensus that offering online access to mental health notes could enhance patients’ understanding about their diagnosis, care plan, and rationale for treatments. These views contrast with findings of small scale surveys in the U.S. and Sweden in which mental health clinicians anticipated most patients would find notes more confusing than helpful.[4,5] There was also consensus that access could enhance patient recall and sense of empowerment about their care plan. The expert panel also agreed that patient access to mental health notes could strengthen multiple relational benefits of care. The panel believed that open notes present no special harms to patients with SMI.

Despite clear consensus on the potential benefits to patient autonomy questions were raised about the effects on professional autonomy. Experts anticipated changes to the detail and accuracy of records with respect to information patients might perceive as negative. Whether the panel believed any such changes might diminish the quality of care, however, was not established. Notably, while experts expect patients to request changes to their notes, they also believed that patient feedback could improve documentation quality. Finally, there was consensus that blocking mental health notes could lead to greater harms including increased feelings of stigmatization. Small scale qualitative studies in psychotherapy suggest that at least some patients experience negative feelings as a consequence of being denied access to their notes.[11,12]

The present study invites a number of important unresolved questions. Perhaps reflecting lack of research on key issues, expert panelists held no strong views about whether patients who disagreed with their diagnosis would be less likely to attend visits or whether too much detail might re-traumatize patients. Our expert panelists also reported “neutral” consensus opinions on whether clinicians would be less detailed/accurate in writing differential diagnoses or in documenting substance abuse disorders. These and other issues, warrant further, more nuanced empirical examination and we strongly recommend that research investigate patients’ experiences with accessing mental health notes across a range of settings, including outpatient and inpatient care, and encompassing a wide range of patient populations with different mental health diagnoses. Focused empirical research is required to better understand whether access influences health outcomes and attendance at visits, and whether patients feel judged or offended by what they read.[25] There is a need for data among patients with serious mental illness, including schizophrenia, personality disorders, or active suicidality to avoid overgeneralizing current evidence to these populations and use cases. Larger scale studies are needed to examine psychiatric clinicians’ experiences including emergent concerns with the practice, and potential sources of patient-clinician disagreement. Finally, it will be important to determine objective changes to documentation as a result of patient access. Combined, empirical research should aim to inform how the innovation might be optimized for its new dual purpose – both as a detailed aide memoire for clinicians and as a tool for communication with patients.

### Limitations

This study has several limitations many of which are inherent to the Delphi methodology. As with all Delphi polls, there are no standard guidelines for identifying expertise. Although we selected participants to represent a diverse spectrum comprising healthcare professionals, patients with lived experiences of mental illnesses, and health informatics researchers, the reliability of consensus opinions in this survey is dependent on the specialist knowledge and experiences of those who participated. While our expert panel represented a diversity of expertise from countries and health organizations where patients are currently offered access to their notes, it is likely that the survey would have benefited from greater ethnic and socio-economic diversity. Notably, challenges related to portal access, and the digital divide, received less attention, and this may have been a consequence of the demographic composition of panelists. Despite these limitations, the study benefited from high response rates between each round.

## Conclusions

In the era of health information transparency, both patient and professional autonomy in mental health contexts must be balanced with the potential benefits and risks to patient care. Experts in this Delphi poll anticipated multiple benefits and few harms of patient access to mental health notes. Further empirical inquiry is required to explore the impact on both patients and clinicians of psychiatric patients of reading their notes, and how both patients and clinicians can become better prepared and supported for documentation transparency in mental healthcare.

## Data Availability

Data available upon request.

## Acknowledgements

The authors express their gratitude to the experts who participated in this survey.

